# The development of IeDEA’s Treat All Dashboard on HIV care outcomes at participating clinics in Sub-Saharan Africa

**DOI:** 10.1101/2025.11.20.25340413

**Authors:** Ellen Brazier, Amanda Berry, Benjamin Katz, Judith T Lewis, Stephany N Duda, Aggrey Semeere, Jacqueline Huwa, Antoine Jaquet, Christella Twizere, Lameck Oteko Diero, Idiovino Rafael, Francois Dabis, Benjamin Muhoza, Denis Nash, IeDEA consortium

## Abstract

Data dashboards are popular tools for communicating information and monitoring progress towards public health goals. To facilitate access to information on key metrics and emergent trends related to the roll-out of universal HIV treatment across diverse settings, the International epidemiology Databases to Evaluate AIDS (IeDEA) leveraged real-world service delivery data from clinics in sub-Saharan Africa to develop 1) a publicly accessible interactive dashboard displaying regional and country-level metrics related to HIV care outcomes across IeDEA sites in sub-Saharan Africa; and 2) a password-protected dashboard displaying similar metrics at the clinic level for staff at sites that contribute data to the IeDEA consortium. This paper describes the development of these dashboards and lessons learned.

## 1. Introduction

The past two decades have brought marked progress towards ending the HIV epidemic, with the estimated number of new infections in 2022 almost halved, and the annual number of AIDS-related deaths decreasing from 1.3 million in 2010 to 630,000 in 2022 [1]. Access to treatment has also improved dramatically with the progressive adoption of the World Health Organization’s (WHO) 2015 recommendation for universal treatment of all persons with HIV (known as “Treat All”), regardless of CD4 cell count or clinical stage; globally, an estimated 76% of those who knew their HIV status in 2022 were on antiretroviral therapy (ART), compared with 48% in 2015 [1].

While nearly all countries in sub-Saharan Africa (SSA) had adopted the WHO 2015 recommendation by 2020 [2], the achievement of global targets, such as UNAIDS’ Fast Track 95-95-95 goals [3], requires up-to-date information of who is being reached through current strategies and programs as well as gaps in the HIV care continuum. To help address this gap, the International epidemiology Databases to Evaluate AIDS (IeDEA) leveraged real-world care data from HIV clinics for web-based visualizations of key metrics and emergent trends in sub-Saharan Africa (SSA)—a region with approximately 660,000 new infections per year and two-thirds of the world’s 39.1 million people living with HIV (PLWH) in 2022 [1]. IeDEA is a global research consortium [4] that curates longitudinal data from HIV clinics in 44 countries across seven regions for HIV-related research. In SSA, more than 200 HIV clinics and programs across 21 countries have contributed data to IeDEA for approximately 1.4 million patients ever enrolling in HIV care [5].

This paper describes the sequential development of two web-based interactive dashboards using data from IeDEA’s patient-level databases in SSA to make trends related to HIV care more accessible to partners and stakeholders at regional, country, and clinic levels: 1) a publicly accessible dashboard displaying regional and country-level metrics related to HIV care and treatment across IeDEA sites in SSA; and 2) a password-protected version with clinic-level metrics for IeDEA sites in Central Africa.

## 2. Methods

We surveyed existing dashboard models [6–8] and key indicators for tracking the global HIV response to identify metrics related to the HIV care continuum that could be derived from aggregations of routine clinical data and developed an initial set of interactive data visualizations, housed on a password-protected site during the development and testing phase. We shared access credentials shared with IeDEA investigators and data managers, and requested their feedback on the content, metrics, and data visualizations through both meetings and an anonymous web-based survey distributed to all users of the test site. Based on user feedback, we refined data visualizations and documentation before making the website publicly accessible. We then undertook a similar development, testing, and review process for the clinic-level version, restricting access to IeDEA partners at contributing sites in Central Africa. We held webinars for in-country partners and circulated a video tutorial and a user guide to familiarize clinic staff and stakeholders with the dashboard metrics and navigation before soliciting feedback through an anonymous web-based survey and direct communication.

## 3. Results

### 3.1. Dashboard Metrics and Data Visualizations

The initial Treat All metrics focused on treatment-naïve patients enrolling in care and initiating ART at participating clinics, including annual enrollments, CD4 count monitoring and CD4 count values at HIV care enrollment, timely initiation of ART, and viral load monitoring and viral load suppression. To allow for faster generation and display of web-based data visualizations and avoid data privacy measures to protect individual-level data, we developed data aggregation scripts that were integrated into IeDEA’s Harmonist Toolkit, a web-based platform for the harmonization and sharing of data across the IeDEA collaboration [9]. Using these scripts, patient-level data for 2010-2020 were aggregated across IeDEA’s four African regions (East, Central, Southern and West Africa), as well as by country, sex, and age-group. After aggregated datasets were prepared, a set of data visualizations were designed and developed for display on a WordPress platform, using Google Charts for visualizations and a MySQL database. The generation of visualizations from aggregated datasets enabled users to apply filters without delayed processing times.

### 3.2. Feedback, Revision and Public Release of the Treat All Dashboard

Feedback from 25 IeDEA investigators and stakeholders informed a series of refinements, including new metrics and data visualizations related to viral load monitoring and suppression, consolidated age groups for data aggregations to minimize small cell counts, small cell suppression for metrics with <20 observations, confidence bands and pop-up messages for improved transparency about uncertainty surrounding estimates, information about unique attributes of selected IeDEA country cohorts, and detailed documentation of the dashboard’s measures and methodology.

After an 18-month development process, the public-facing IeDEA Treat All Dashboard (iedeadashboard.org)—with interactive data visualizations on key metrics of interest for 2010 through 2017 (**Figure 1**)—launched in September 2021 with a French-language version launched in February 2022. To contextualize IeDEA data, population-based information about the number of new HIV infections and the number of people with HIV was compiled from UNAIDS data for each year and country. Once launched, the Treat All Dashboard was promoted through regular IeDEA communications (e.g., newsletters, Twitter/X) to researchers, policymakers, and other stakeholders within and beyond IeDEA. The website was visited by more than 500 users in 27 countries within the first four months of being launched.

**Figure 1.**
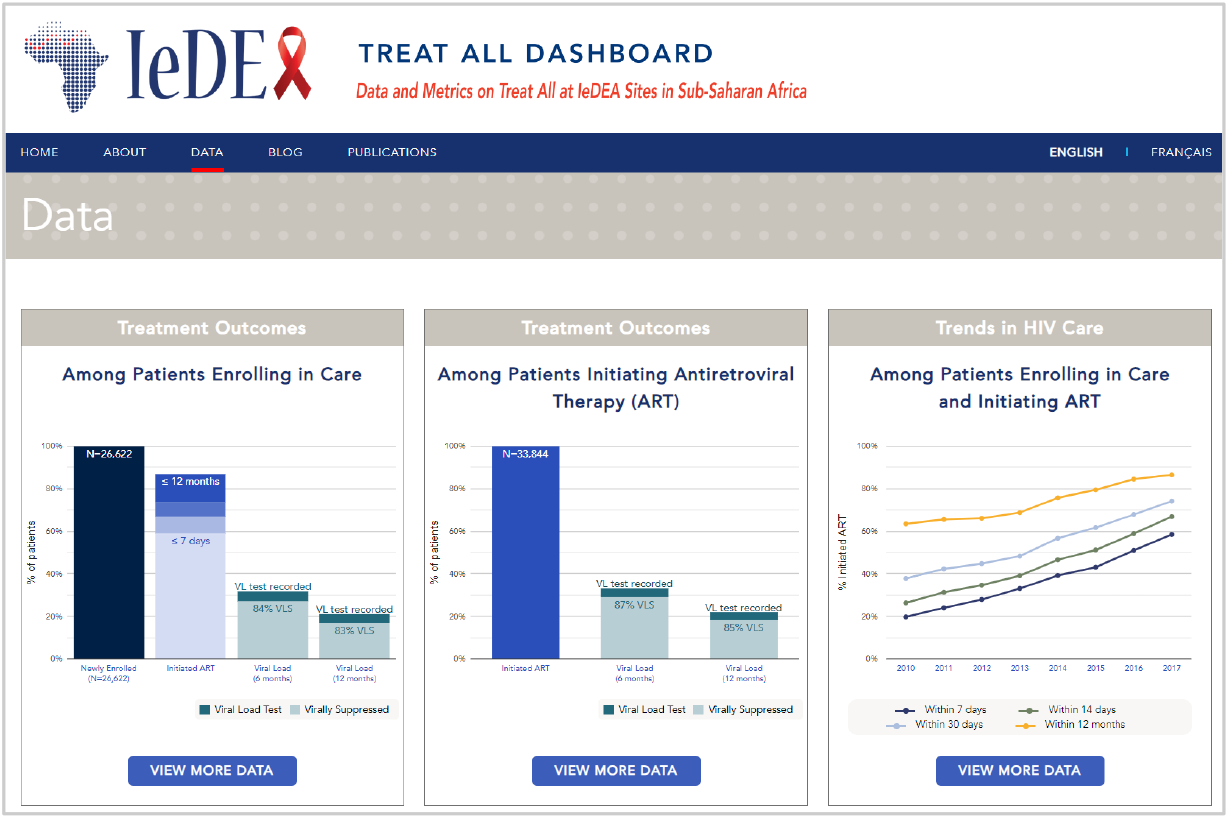
Screenshot of the Treat All Dashboard showing interactive data visualizations

### 3.3. Adaptation of Treat All Dashboard for IeDEA clinics

For the clinic-level Dashboard created for IeDEA partners at 21 contributing sites in Central Africa, data suppression rules were removed to enable clinic staff to view counts and trends for all patients included in Dashboard visualizations, irrespective of small cell counts. Clinic users could view metrics for their clinic only, for their clinic alongside aggregated data for all IeDEA clinics in the same country or all IeDEA clinics in SSA.

### 3.4. Feedback from Clinic Users

Feedback on the clinic-level Dashboard from 23 clinical and administrative staff across 17 clinics in five countries was overwhelmingly positive, with approximately 90% reporting that it was easy to access their clinic’s data and 83% indicating data visualizations would be useful for their work. However, in response to open-ended questions about potential uses of the Dashboard, most respondents outlined uses that centered around real-time individual patient monitoring to improve treatment adherence, care retention, and viral suppression and reporting—uses that would require identifiable individual-level data. Feedback suggested that global programmatic metrics and targets used to measure progress towards ending the HIV epidemic (e.g. timely ART initiation), may be less salient for clinicians charged with monitoring patients’ health and well-being.

## 4. Discussion

IeDEA’s public-facing Treat All Dashboard was well received and initially visited by large numbers of users. User traffic decreased during subsequent months, however, underscoring the need for timely updates to maintain user engagement, as well as the declining interest in Treat All once most countries around the globe had adopted WHO’s universal treatment recommendations. Although regular data updates had been part of the initial Dashboard plan, inherent lags of 18-24 months in receiving and processing data from IeDEA sites constrained rapid updates. Additionally, high levels of customized programming used to contextualize Treat All trends and metrics and ensure transparency about small cell counts made updating the visualizations time and resource intensive.

New initiatives launched by the global IeDEA consortium also reduced the need for regular Dashboard updates. To improve the accessibility of the consortium’s data to researchers and decision-makers, IeDEA began working to develop a shareable “multiuse dataset” with associated web-based visualizations, which includes data on regions outside of SSA and time periods beyond Treat All implementation. IeDEA’s multiuse dataset development was informed by the Treat All Dashboard experience, with developers enforcing a strict data preparation pipeline and limiting user-requested customizations to improve timeliness of data updates.

For the password-protected, clinic-level version of the Dashboard, the removal of data suppression requirements required less customized programming and allowed for simpler refreshes with updated data, but the use of aggregated data precluded the type of individual-level patient tracking of primary interest to clinician partners at Central Africa IeDEA sites. The Dashboard’s focus on Treat All trends and HIV care cascades for patients was not well aligned with the day-to-day priorities of staff involved in patient care and reporting—both of which require real-time data on a clinic’s active patient population, as opposed to aggregated metrics. In the future, integrating HTML/CSS-based visualizations into routine processing of clinic data may be a more efficient way to provide timely updates and visualizations of interest to clinic partners.

## 5. Conclusion

While initially successful in catalyzing engagement with IeDEA data related to Treat All metrics, the development of IeDEA’s public-facing and clinic-level Dashboards underscored the fact that such data dashboards may inevitably become less relevant as programmatic priorities evolve over time. Additionally, while it is critical to develop tools that enable and encourage clinic-level staff to access and engage with their data, the metrics most relevant to clinician partners may differ substantially from those of interest to researchers and program planners. Addressing the needs of clinicians may require more complex data processing and data privacy elements than is required for visualizations of aggregated data related to programmatic goals and targets. Understanding the specific needs of end users and ensuring the capacity for rapid data refreshes by minimizing highly customized data visualizations are critical for producing data dashboards with lasting value and relevance.

## Data Availability

The IeDEA Treat All Dashboard is a public website

https://iedeadashboard.org/

## Acknowledgements

The International Epidemiology Databases to Evaluate AIDS (IeDEA) is supported by the U.S. National Institutes of Health’s National Institute of Allergy and Infectious Diseases, the *Eunice Kennedy Shriver* National Institute of Child Health and Human Development, the National Cancer Institute, the National Institute of Mental Health, the National Institute on Drug Abuse, the National Heart, Lung, and Blood Institute, the National Institute on Alcohol Abuse and Alcoholism, the National Institute of Diabetes and Digestive and Kidney Diseases, and the Fogarty International Center: Central Africa, U01AI096299; East Africa, U01AI069911; Southern Africa, U01AI069924; West Africa, U01AI069919. Informatics resources are supported by the Harmonist project, R24AI24872. This work is solely the responsibility of the authors and does not necessarily represent the official views of any of the institutions mentioned above. This work is subject to the NIH Public Access Policy. Through acceptance of this federal funding, NIH has been given a right to make this work publicly available in PubMed Central upon the Official Date of Publication, as defined by NIH.

